# Identifying the Presence and Timing of Self-harm in Electronic Mental Health Records Using Privacy-Preserving Local Language Models: Methodological Study

**DOI:** 10.1101/2025.10.27.25338892

**Authors:** Andrey Kormilitzin, Dan W. Joyce, Apostolos Tsiachristas, Rohan Borschmann, Navneet Kapur, Galit Geulayov

## Abstract

**Background:** Self-harm is the strongest risk factor for suicide and an important outcome for mental health care. Although prevalent in clinical populations, it is often imprecisely captured in routinely collected clinical data, where it is often recorded and stored as unstructured free text. Contemporary language models, such as GPT (OpenAI) and Gemini (Google), can analyse free-text clinical notes, but such cloud-based commercial and closed-source models may violate data governance of processing sensitive patient data.

**Objectives:** We evaluated whether a privacy-preserving language model running entirely within an institution’s secure computing infrastructure (here, the UK National Health Service; NHS) could accurately identify the presence and timing of self-harm using electronic health records (EHRs) from secondary mental healthcare.

**Methods:** Clinical notes were drawn from Oxford Health NHS Foundation Trust using a multi-stage workflow: (1) a random sample of 1,000 patients with a psychiatric diagnosis (ICD-10 F00–F99); (2) candidate-note identification using a Gemma3-4b language model to flag notes containing self-harm content; (3) from those candidates, 1,352 randomly sampled notes were selected for expert annotation. The resulting gold-standard corpus is therefore enriched for self-harm content. Each clinical note was annotated for the presence/absence of self-harm and its timing (≤90 days/>90 days/unknown). A privacy-preserving locally served 27-billion-parameter Gemma3 language model (’Gemma3-27b’) was used as the core model. Prompts were systematically developed and refined using a labelled development set to identify self-harm and generate a structured output per clinical record. The performance of Gemma3-27b model was compared against a strong baseline multi-label text classification model based on RoBERTa (Robustly Optimized BERT Pretraining Approach, a transformer-based language model) architecture. Model performance was evaluated using precision, recall, and the F1-score (harmonic mean of precision and recall), with 95% confidence intervals estimated from 1,000 bootstrap samples with replacement.

**Results:** Gemma3-27b outperformed the RoBERTa classifier across all categories, achieving Precision=0.92, Recall=0.92, and F1-score=0.92 for notes containing self-harm, and Precision=0.97, Recall=0.97, and F1-score=0.97 for notes without self-harm. For the 51 notes labelled as recent self-harm in the held-out test set, Gemma3-27b achieved Precision=0.84, Recall=0.75, and F1-score=0.79. The global weighted F1-score of Gemma3-27b across all categories was 0.88, compared to 0.85 for RoBERTa.

**Conclusions:** With systematic prompt development on a labelled development set, but no gradient-based fine-tuning, the current Gemma3-27b language model matched or exceeded a fine-tuned RoBERTa classifier for ascertaining self-harm events and their timing. Aggregate gains were modest, while improvements were largest in the most challenging, lower-frequency timing categories. On a simplified binary recent-versus-other task, RoBERTa performed marginally better, indicating that supervised classifiers remain highly effective when the task is simplified and sufficient labelled data exist. This work demonstrates the technical feasibility of privacy-preserving self-harm detection within a secure NHS research environment.

## Introduction

Self-harm (intentional self-poisoning or self-injury, irrespective of motivation [1]) represents a major public health challenge. In England, approximately 5,000 individuals die by suicide each year [2] and more than 200,000 individuals present to general hospitals due to self-harm [3]. Many more self-harm without seeking treatment [4].

Self-harm is the strongest risk factor for suicide [5]. Individuals who present to clinical services following self-harm are over 100 times more likely to die by suicide compared to those who do not self-harm [6, 7]. Their risk of accidental death and death by natural causes is also markedly elevated [5]. Furthermore, these individuals have a higher risk of further non-fatal self-harm and adverse psychosocial outcomes [6, 8, 9].

Despite it being an important outcome for mental health care, information about self-harm is often imprecisely captured in many healthcare settings. For example, in one study from England [10], the investigators compared research-derived rates of hospital-presenting self-harm to official Hospital Episode Statistics (HES) data. The study found substantial under-ascertainment in official statistics compared with the research-derived figures, even though both sources drew on the same underlying clinical information. Accurate and systematic identification of self-harm across settings is essential for conducting valid and reliable research and for planning and delivery of effective intervention strategies.

Suicide and self-harm research involves numerous methodological challenges. It can be resource-intensive and costly, with data collected and collated over many years from some (but not all) relevant settings. Consequently, many instances of self-harm go undetected, leading to missed opportunities for intervention and compromised research. Leveraging existing data collected as part of routine patient care can provide a valuable, contemporaneous and economical source of information. Such data, which contain a wealth of information, have been used to study many health conditions e.g. cardiovascular disease, diabetes, and osteoarthritis [11, 12]. However, utilising such rich information comes with significant challenges, particularly due to the large volume of data, much of which is often collected and stored in an unstructured narrative format. Advances in artificial intelligence (AI) and natural language processing (NLP) present an opportunity to unlock, retrieve, and convert this information into a format accessible for research and clinical care.

Previously, investigators have used the Clinical Record Interactive Search (CRIS) database of the South London and Maudsley NHS Foundation Trust to identify suicidal ideation and self-harm from free text in secondary mental health EHRs [13]. Such models show good performance in identifying patients who have self-harmed. Identifying the timing of the self-harm through free text, however, has been more challenging [14]. The timing of self-harm is important for both research and clinical practice. Evaluating the effect of interventions or routine care depends on accurately establishing the timing of self-harm. Similarly, reliable longitudinal analysis relies on ascertaining the temporal sequence of self-harm alongside its covariates. Importantly, the risk of suicide and repeat self-harm is acutely elevated soon after a self-harm episode [7, 15]. As such, accurately capturing the timing of self-harm episodes is critical for identifying individuals in need of timely interventions and risk reduction strategies.

### Machine learning for self-harm identification

Well-established machine learning models for typical NLP tasks such as named-entity recognition, relationship extraction, text classification tasks, and negation detection have shown good ability to identify and structure the concepts of interest [16]. However, training such models relies on a large amount of data, manually annotated by experts. Collecting a sufficient amount of high-quality annotated data can be an expensive and time-consuming task. Since the introduction of large language models (LMs) and, in particular, Generative Pre-training Transformer (GPT) and their chatbot interface such as ChatGPT, the information extraction field has seen a paradigm shift. Multiple studies have shown that generic large LLMs (e.g., GPT, Claude, Gemini) trained on a massive corpus from the internet, can identify concepts of interest and generate a structured output following the prompt tailored for each particular task [17]. For instance, LLMs have been successfully applied to extract complex relationships between biomedical entities from the scientific literature by carefully prompting the model with a description of the desired relationship [18]. Furthermore, related prompt-or question-answering formulations have also been explored for event extraction (e.g., machine reading comprehension) and improved performance in data-scarce settings [19]. Additionally, recent work has explored the use of LLMs for computational social science tasks involving noisy and ambiguous user-generated text, including social media data [20].

However, the use of proprietary LM services via their application programming interface (API), such as those provided by OpenAI (GPT), Anthropic (Claude) and Google (Gemini), poses significant challenges to patient data privacy and may not be compliant with clinical information governance. In contrast, if a language model can be implemented within the healthcare provider’s own secure clinical data environment there is no need to risk exposing sensitive and confidential data via APIs to proprietary services. Until recently, implementing LLMs (including training and inference) has been implausible because of their memory and computing costs. With the introduction of quantised LLMs, models that have been made smaller and more computationally efficient by storing the numerical values of their parameters in a simpler form, open-weight models such as Gemma3-27b can be hosted and used for inference on modest compute resource with performance (for specific tasks) only marginally lower than the original (not quantised) model. Therefore, researchers have explored the use of these local versions of LLMs, such as Llama [21] for information extraction from clinical records [22, 23], as well as for identifying acts of suicidality [24].

### Motivation and our contribution

In this study we evaluated whether privacy-preserving local language models can identify self-harm and its timing in secondary mental health records, converting unstructured clinical notes into structured data. We assessed the semantic reasoning capabilities of a pre-trained language model to distinguish self-harm events from related concepts (e.g., ideation, risk assessments) and classify their timing. Specifically, we tested whether an open-weight Gemma3 model with 27 billion parameters, deployed locally, can accurately detect self-harm and identify its timing without gradient-based fine-tuning (i.e., without updating the model’s internal parameters on our data), relying instead on systematic prompt development using a labelled development set.

We compared the current approach against a supervised RoBERTa classifier (a commonly used model) trained on identical data. Model performance was assessed using precision, recall, and the F1-score (the harmonic mean of precision and recall) with 95% confidence intervals estimated using 1,000 bootstrap samples with replacement. We hypothesised that local language models would: (i) match or exceed supervised model performance through prompt-based inference guided by a labelled development set, thereby reducing the volume of annotated data needed for gradient-based training; and (ii) mitigate data governance barriers inherent in using cloud-based solutions via application programming interfaces (APIs), enabling deployment within healthcare institutions.

This work addresses the critical need for accurate self-harm identification in clinical records, using language models that can be deployed locally under strict patient confidentiality standards for sensitive mental health data and within constrained computational resources.

## Methods

### Definition of self-harm

Self-harm refers to any form of intentional self-poisoning or self-injury, irrespective of motivation [1]. It can take many forms, including overdosing on medications, ingesting a non-ingestible substance, or inflicting injury upon oneself through actions like cutting. In clinical settings (e.g., hospital emergency departments and mental health services), self-harm ascertainment relies on clinician’s judgement; i.e., a clinician will determine whether the self-inflicted act was intentional, as opposed to accidental, even in the absence of patient confirmation [25].

### Data source and ethics

Data for this study was sourced from the Clinical Record Interactive Search (CRIS) system by Akrivia Health analytics platform on behalf of the Oxford Health NHS Foundation Trust, UK. Akrivia Health provides a secure research environment with a robust information governance framework compliant with national statutory regulations for healthcare data. The CRIS database comprises pseudonymised EHRs including free-text clinical notes as well as structured data fields from secondary care mental health services [26]. Studies using this platform require approval from the healthcare institution that provided the data. This study received approval from the Oxford Health NHS Foundation Trust CRIS Oversight Committee and data were processed in accordance with the procedure outlined by the Oxford Health NHS Foundation Trust [27].

### Cohort selection

The study population involved individuals aged 18 years or over with a confirmed psychiatric diagnosis (see Appendix A) according to the International Classification of Diseases, Tenth Revision (ICD-10), who were in contact with specialist secondary mental healthcare services. Clinical records of patients with primary diagnoses of ICD-10 codes F00-F99: Mental and Behavioural Disorders (see Appendix A), recorded between 1 March 2016 and 1 March 2022 (inclusive) were randomly sampled for annotation, as described in Figure 1.

**Figure 1:**
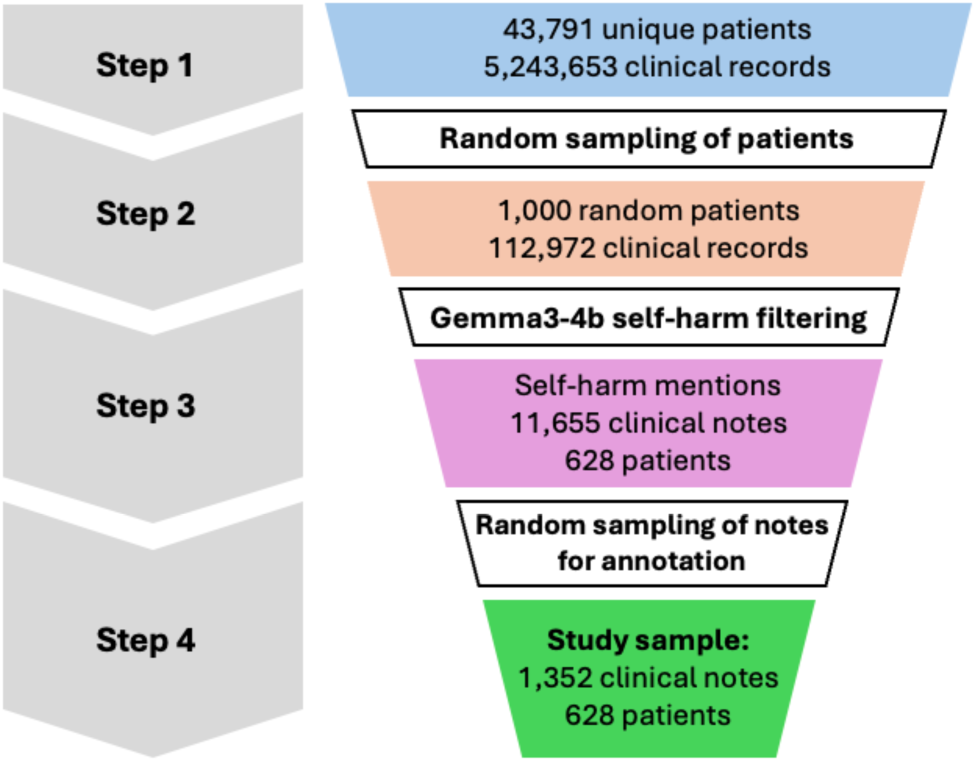
The data flow diagram of the clinical notes used to develop the models.

In contrast to a common approach of identifying clinical notes that contain mentions of self-harm using a keyword-based fuzzy pattern matching (which can be biased in identifying cases beyond the predefined keywords), we opted for using a small, yet capable LM (‘Gemma3’) with four billion parameters (‘Gemma3-4b’) prompted to identify potential self-harm events minimising the risk of the aforementioned keyword-based bias (Appendix B).

### Data preparation workflow

Figure 1 provides an overview of the end-to-end data selection process in this study. First, an initial cohort of 43,791 patients with over 5.2 million clinical notes satisfying the above criteria, was extracted from the Oxford Health NHS CRIS database (Step 1). Given the limited human annotator resources, we randomly sampled 1,000 patients (Step 2), followed by retrieving clinical notes containing potential mentions of self-harm using a lightweight “Gemma3-4b” LM (Step 3). However, the model identified over 11,000 notes containing self-harm, which significantly exceeded the capacity of our clinical annotators. As such, we further randomly reduced the cohort of 11,655 clinical notes containing self-harm to 1,352 clinical notes from 628 unique patients (Step 4).

Since we used the Gemma3-4b language model to initially identify clinical notes that might contain self-harm cases, the text we found was richer in this type of content than one would find across all patient records. To mitigate this selection bias, we further randomly sampled 1,352 notes from 628 different patients for expert review. While our annotated dataset likely contains more self-harm language than a typical sample would, the random selection process made sure we weren’t inadvertently favouring particular patients, specific time frames, or certain diagnostic groups.

This enrichment improves annotation efficiency by increasing the proportion of positive examples available for model development, but it changes the class distribution relative to routine care. In particular, model performance metrics, e.g., positive predictive value (PPV) may differ when applied to all clinical notes, where the prevalence of self-harm mentions is substantially lower. Implications for routine deployment are discussed in the Limitations section.

### An annotation schema and a curated dataset

Manual annotation of textual data is essential for developing and evaluating NLP models for information extraction. A systematic annotation process involving expert coders ensures unambiguous tagging of text segments according to a predefined schema, enabling models to learn meaningful patterns and providing a human benchmark for performance.

We used a multi-label annotation schema focusing on whether an actual act of self-harm occurred and whether it was i) recent (occurring within 90 days of documentation), ii) historical (those occurring >90 days prior to documentation), or iii) of unknown timing (the timing could not be determined). Specifically, if annotators identified a self-harm event, the corresponding text was labelled as “Self-harm present” along with a timing tag. Where it was not possible to unambiguously determine whether a self-harm event took place, we labelled these cases as “unknown self-harm”. Clinical notes that did not include self-harm events (e.g., a clinical note describing a patient with psychotic symptoms with no mentions of self-harm or one that mentions risk of self-harm but not actual self-harm) were unlabelled. The annotation protocol included initial training and calibration sessions, detailed guidelines with clinical examples, and regular inter-rater reliability assessments. Disagreements were jointly reviewed to refine decision boundaries and for consistency. The schema thus comprises five distinct labels as shown in Table 1.

**Table 1.** Five categories to designate the self-harm events used in the study.

| Label | Explanation |
| --- | --- |
| Self-harm absent | Statement negating self-harm or where self-harm was not mentioned at all (e.g., a clinical note recording only psychotic symptoms) |
| Self-harm present | Explicit description of a self-harm (e.g., self-poisoning or self-injury) event by the patient. |
| Recent | Event occurred within the last 90 days prior to the note date |
| Historical | Event occurred more than 90 days prior to the note date |
| Unknown timing | Where time cannot be determined from the information provided (e.g., “the patient has self-harmed previously” or “Deliberate self-harm scars and burns evident”). |

All forms of intentional self-inflicted harm (including suicide attempts and self-harm where the specific motivation was not explicitly mentioned) were in scope; self-harm ideation (e.g., “patient feels like cutting”, “patient wishes to end it all”) were excluded unless a self-harm act was also mentioned.

Many patients had a long-documented history of contact with secondary mental health services. Annotators were instructed to treat each clinical note extract as a standalone document, independent of any decisions made about previous extracts from the same patient.

### Partitioning of data for training and validation

The annotated sample of 1,352 examples was split at the patient level, to avoid data leakage, into the training (80%, n=1084) and test (20%, n=268) sets, respectively. Resulting class distribution within the training and test splits is summarised in Figure 2. In most instances, when self-harm was mentioned by a clinician, there was sufficient information to determine whether this was a recent or past event.

**Figure 2:**
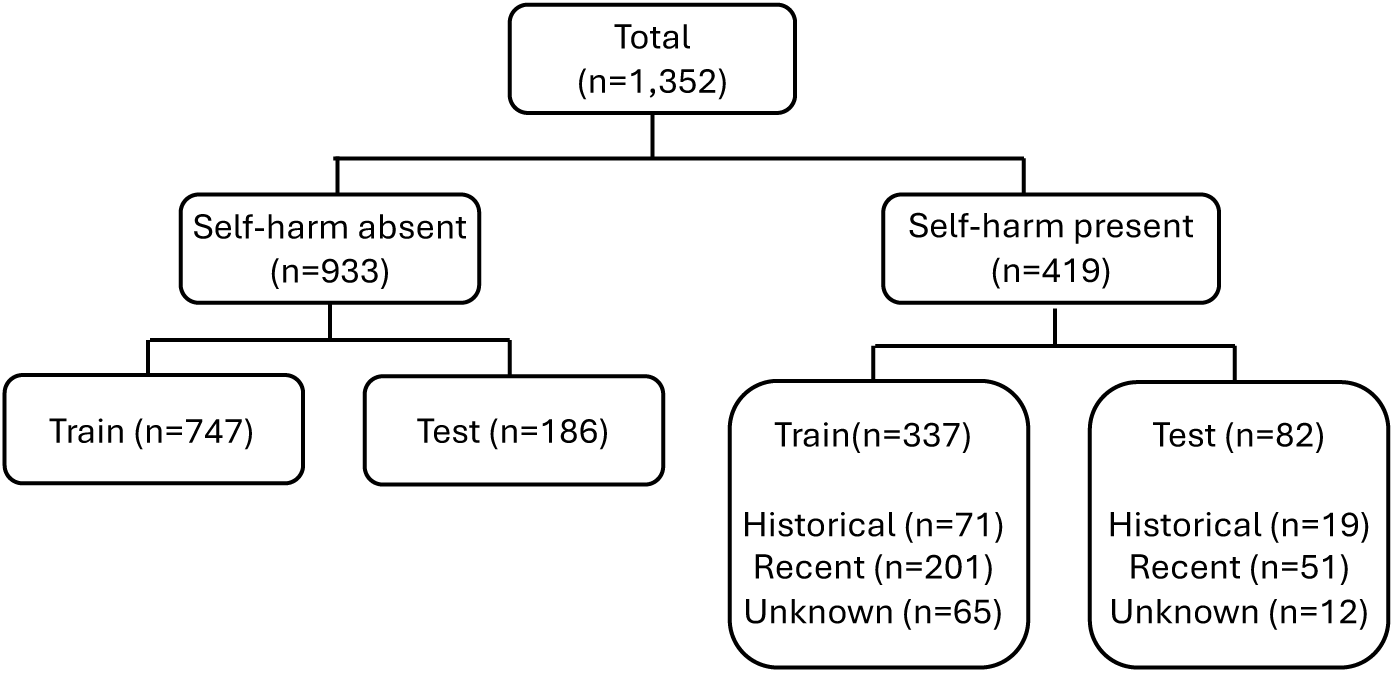
The distribution of 1,352 clinical notes in the gold standard corpus according to self-harm status and timing labels, by training (80%) and testing (20%) sets.

### Annotation procedure

In order to evaluate the developed annotation schema and the degree of agreement, two data annotators, including one researcher (LB) with over 30 years’ experience collecting and coding self-harm information from hospital records in the UK, and another (GG) with more than 15 years’ experience researching self-harm and suicidal behaviour, refined category boundaries and created a decision flowchart. They subsequently independently further labelled 1,352 randomly sampled notes.

### Comparing the agreement on annotations

Inter-annotator agreement (IAA) was calculated on a sample of 160 notes annotated independently by two raters, using a Cohen’s 𝜅 within a hierarchical evaluation framework. Each clinical note was annotated along two categories: 1) self-harm status (“present” or “absent”) and 2) timing of self-harm (“recent”, “historical”, or “unknown”). The timing labels were sought only if self-harm was “present”. As the second decision category (i.e., timing of self-harm) was conditional on the first category, we opted to report IAA using a hierarchical protocol: i) assess agreement on self-harm status (present/absent); ii) assess agreement on the timing of self-harm event (“recent”, ”historical”, or “unknown”) within the subset of notes where both annotators agreed that self-harm was present.

For category one - self-harm present or absent - each note contributed to a 2-by-2 contingency table and the resulting 𝜅*_self-harm_* captured chance-corrected concordance on event detection. Using the subset of notes where both coders marked self-harm present, each annotator assigned one of three nominal categories: recent, historical or unknown, yielding a 3-by-3 table from which an unweighted 𝜅*_recency_* was computed. The 3-by-3 structure reflects the three possible timing categories assigned independently by each of the two annotators. Uncertainty estimates for both *IZ* coefficients were obtained via 1,000-fold stratified bootstrap resampling with replacement from the 160 jointly annotated notes. At each iteration, the set of notes was resampled while preserving the marginal class distribution (i.e., the proportion of “present” vs “absent” for self-harm status, and the relative frequencies of “recent”, “historical”, and “unknown” within the subset marked as self-harm present). For each bootstrap replicate, 𝜅 was recalculated, and the 2.5th and 97.5th percentiles of the resulting empirical distribution were used to form 95% confidence intervals. This stratification ensured that class imbalance did not distort the variability estimates of *IZ*.

### Model development Core language model

While language models have demonstrated strong capabilities in clinical text processing and reasoning, their computational demands can be prohibitive [28]. Therefore, we explored the Gemma3-27b, a decoder-only transformer model with 27 billion parameters and a 128K token context window [29]. The model is based on a novel architecture with a 5:1 ratio of local to global attention layers, where local layers employ sliding window attention over 1,024 tokens to reduce memory consumption during inference. The model was quantised to 4-bit precision (Q4_K_M format) resulting in a 10.6 GB model size and served locally within a secure environment at the Oxford Health NHS Foundation Trust using the Ollama framework (v0.9.6) with llama.cpp backend [30]. All experiments were conducted on the Microsoft Azure T4 instance (‘Standard_NC8as_T4_v3’) with 16GB GPU memory as a cost-effective solution for the NHS for information extraction and reasoning tasks.

### Baseline multi-label text classification

To evaluate the benefit of language models for semantic reasoning on self-harm, we trained and evaluated a transformers-based text classification model as a baseline (a benchmark model). We chose the RoBERTa model for its competing overall performance and speed. The model was fine-tuned using a binary cross-entropy (BCE) with logits loss, natively implemented in PyTorch [31]. The reason for opting for a multi-label text classification model was to mimic the behaviour of a language model, whereby it outputs simultaneously both self-harm status and timing labels. For consistency, the RoBERTa model was trained and evaluated using the same training and test data splits used to develop the Gemma3 model. For reproducibility and detailed training, see Appendix B.

### Prompt engineering

The effective use of language models for information extraction relies heavily on well-designed prompts that provide clear instructions and context for the task at hand. In this study, we focused on specifying rules and contextual cues to identify self-harm and determine its timing. Furthermore, our prompt design addressed the challenge of distinguishing actual self-harm events from related concepts such as suicidal ideation and self-harm risk. The prompt development followed established prompt-engineering principles, including prompt programming, prompt-based learning and chain-of-thought-style task decomposition [32,33,34], to address two sequential classification tasks: (i) binary detection of self-harm presence, and (ii) temporal classification into three categories.

The prompt design incorporated clear inclusion criteria for completed intentional acts, comprehensive exclusion criteria (e.g., thoughts, plans, threats), and specific guidance for ambiguous cases. The final prompt required JSON-formatted output with direct textual evidence for each classification, using a 90-day threshold for determining recent episode of self-harm. The prompt was validated by two self-harm experts before deployment.

To mitigate the risk of prompt overfitting and to meaningfully compare to a baseline RoBERTa model, the models were trained and tested on the same split partitions (Figure 2). For the Gemma3-27b iterative prompt refinement procedure, the training data set was further split into development (n=542) and validation (n=542) datasets. The optimal prompt was developed iteratively on the development set (n=542), and after satisfactory performance was achieved on the validation set (n=542), the model was finally evaluated on the held-out test set (n=268).

For RoBERTa, we trained two variants: RoBERTa (n=542) using only the development set for fair comparison with Gemma3-27b’s prompt refinement data exposure, and RoBERTa (n=1,084) using the full training set. Both models were evaluated on the identical held-out test set, ensuring unbiased performance comparison. The data flow through our experimental pipeline is illustrated in Figure 1. The prompt engineering approach with all development details and the prompt used in this work are presented in Appendix C.

## Results

### Annotation consistency

The results of inter-annotator agreement are shown in Table 2. These include confidence intervals calculated using 1,000 bootstrap resamples on a random set of 160 notes annotated independently by two human experts. The agreement was *very good* for self-harm status (presence/absence) and was *good* for timing on self-harm-present notes [35], indicating robust and unambiguous annotation rules and the ability of two independent annotators to follow it easily.

**Table 2.**
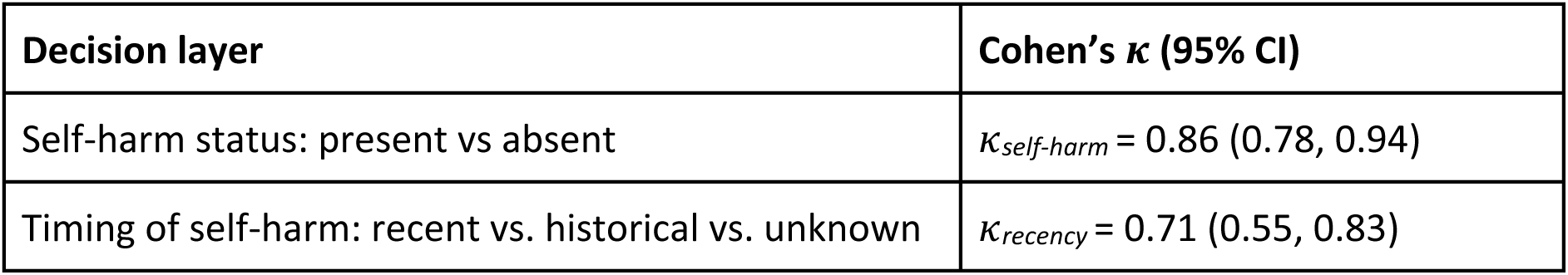
Inter-annotator agreement for identifying self-harm and its timing.

| Decision layer | Cohen’s $\kappa$ (95% CI) |
| --- | --- |
| Self-harm status: present vs absent | $\kappa_{self-harm} = 0.86$ (0.78, 0.94) |
| Timing of self-harm: recent vs. historical vs. unknown | $\kappa_{recency} = 0.71$ (0.55, 0.83) |

### Multi-label self-harm identification models

Unless otherwise specified, we report F1-score. Sensitivity, specificity, and recall metrics are reported explicitly. Although Gemma3-27b requires no task-specific fine-tuning, we nevertheless built two supervised baselines to benchmark its zero-shot prompt-based extraction. The first, RoBERTa (n=542), was trained on the same 542-note development split that guided prompt refinement, giving a like-for-like comparison in terms of labelled data “seen” by each approach. To assess the models’ performance, we computed the point estimates for precision, recall and F1 score for each of the five categories: (1) at the self-harm status level: present and absent, and; (2) at the timing level: recent self-harm episode, historical episode, and unknown timing. The performance metrics with corresponding 95% confidence intervals are presented in Table 3.

**Table 3:**
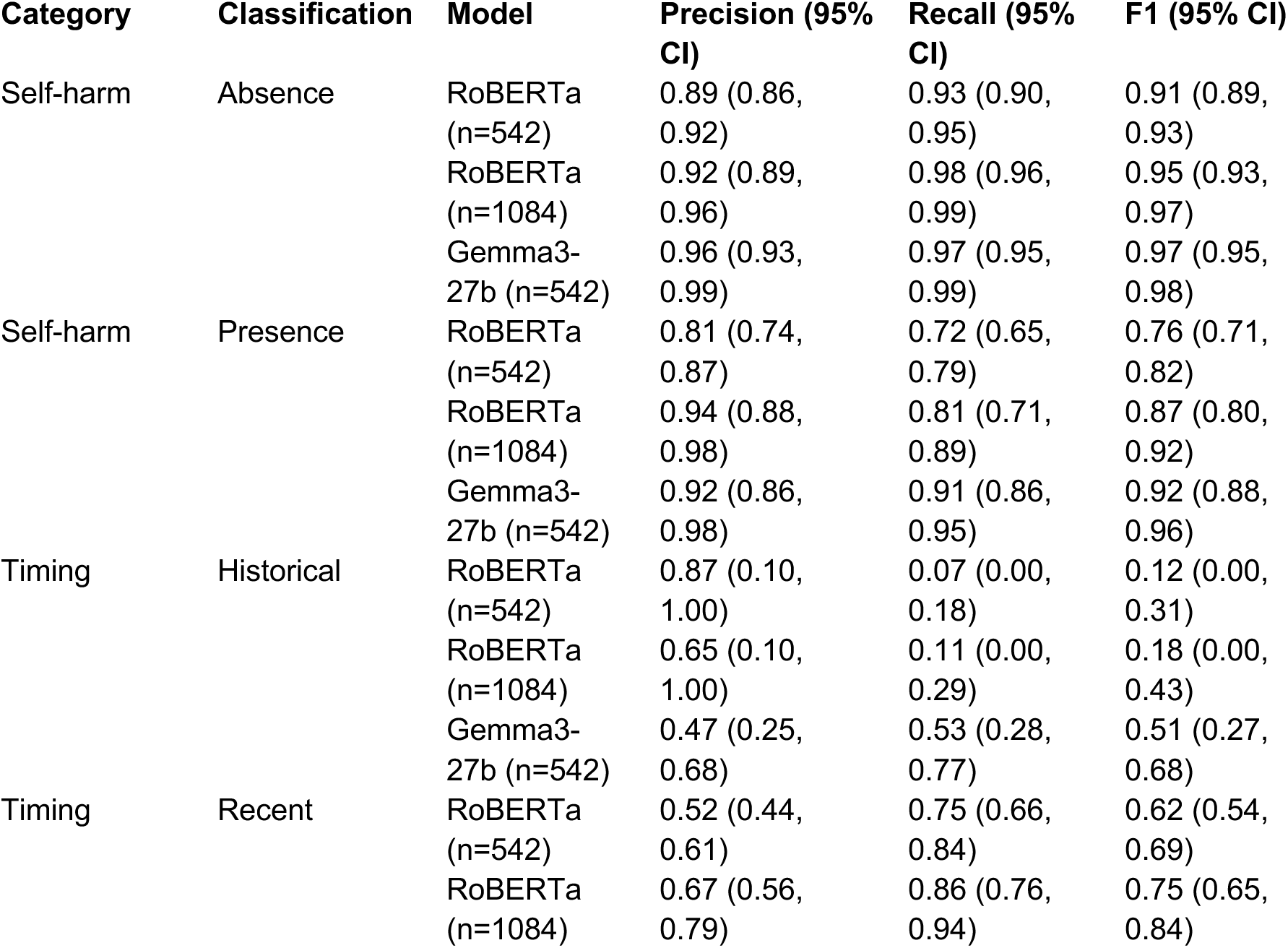
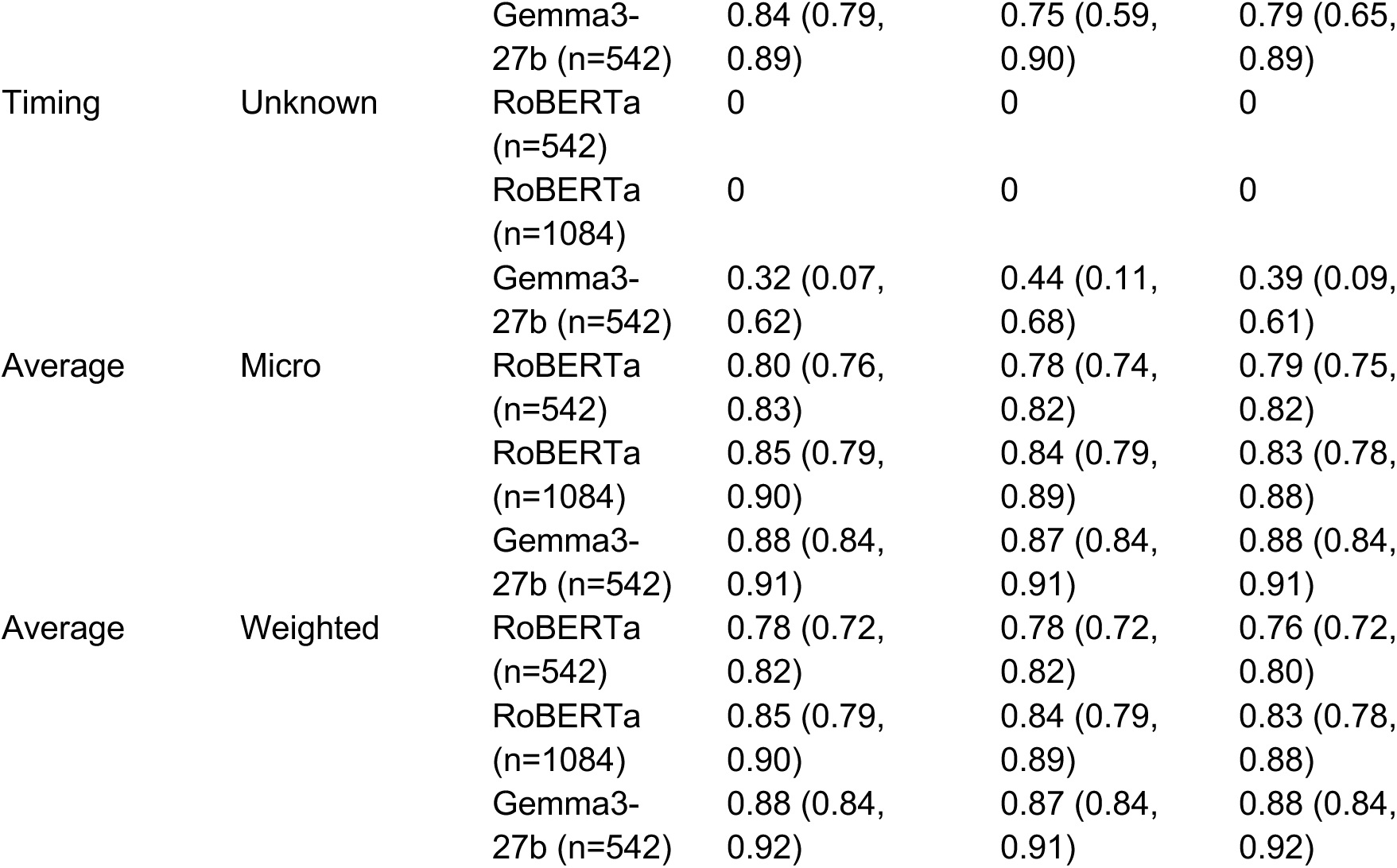
Comparisons of the three models’ performance according to self-harm status and its timing labels. Two baseline RoBERTa models were trained on datasets with 542 and 1084 samples

Table 3 shows that RoBERTa (n=542) underperformed Gemma3-27b across all labels, most notably on recent self-harm (F1 = 0.62 vs 0.79) and on the low prevalence historical class, where recall collapsed to only 7% vs 53% that of Gemma3-27b. The multi-label confusion matrices for all categories are shown in Figure D.1 in Appendix D.

We assumed that weak performance of RoBERTa could be attributed to limited training examples rather than intrinsic model capacity. Accordingly, we trained a second baseline RoBERTa (n=1,084) on the full 1,084-note training set. Performance improved, but even with nearly double the annotations RoBERTa still failed to surpass Gemma’s precision-recall balance, demonstrating that even a relatively small (27 billion parameters) privacy-preserving language model offers a stronger starting point than a task-specific transformer, even when the latter is given all available data.

The Gemma3-27b model demonstrated superior performance across all classification categories, achieving a weighted F1-score of 0.88 and micro F1-score of 0.88, compared to RoBERTa’s 0.83 and 0.83, respectively. This is consistent with the broader pre-trained knowledge that large language models bring to semantically complex clinical tasks, although the aggregate gains were modest (≈ 3-5 weighted F1 points). The disparity was particularly pronounced for the more challenging temporal categories. RoBERTa failed entirely to identify “unknown timing” cases (F1=0.0) and performed poorly on “historical” classifications (F1=0.18), while Gemma3-27b achieved F1-scores of 0.39 and 0.51 for these categories, respectively. This stark difference highlights a fundamental limitation of supervised approaches when training data are scarce. Both categories were rare in the corpus, with 77 of 1,352 notes (5.7%) labelled as “unknown” and 90 of 1,352 notes (6.7%) as “historical.” The performance of two leading models, Gemma3-27b and RoBERTa (n=1084), were compared using McNemar’s test for multi-label classifications with bootstrap analysis and the Benjamini-Hochberg false discovery rate (FDR) method for multiple comparison correction. All details of the statistical comparison are shown in Appendix E.

The RoBERTa model’s difficulty with rare categories illustrates a well-known challenge in clinical NLP: obtaining sufficient annotated examples for every category is often impractical. Supervised learning typically requires many examples per class to achieve reliable performance, a requirement rarely met for infrequent but clinically important categories.

Gemma3-27b’s relatively stronger performance on these rare categories, achieved through iterative prompt engineering on a labelled development set rather than gradient-based training, suggests that the model’s pre-trained knowledge provides a useful starting point for handling the long-tail distribution typical of clinical data. However, we note that this advantage was most evident in lower-frequency timing categories; aggregate gains over RoBERTa trained on the full dataset were modest. For reproducibility and technical details of models training, please refer to Appendix F.

### Binary classification for recent self-harm detection

To evaluate real-world applicability, we reformulated our multi-label task as a binary classification problem focused on identifying recent self-harm - the most clinically actionable category.

This approach mirrors practical use cases where, for example, clinicians may need to identify patients requiring intervention. This approach provides a simplified and practical categorisation, aimed at identifying individuals with a recent self-harm event. We combined the original labels into two categories: (i) “Recent self-harm” - cases with confirmed self-harm occurring within 90 days (n=252), and (ii) “Other events - all remaining cases, including absent self-harm, historical events, or unknown timing (n=1,100), as shown in the data flow chart in Figure 3.

**Figure 3:**
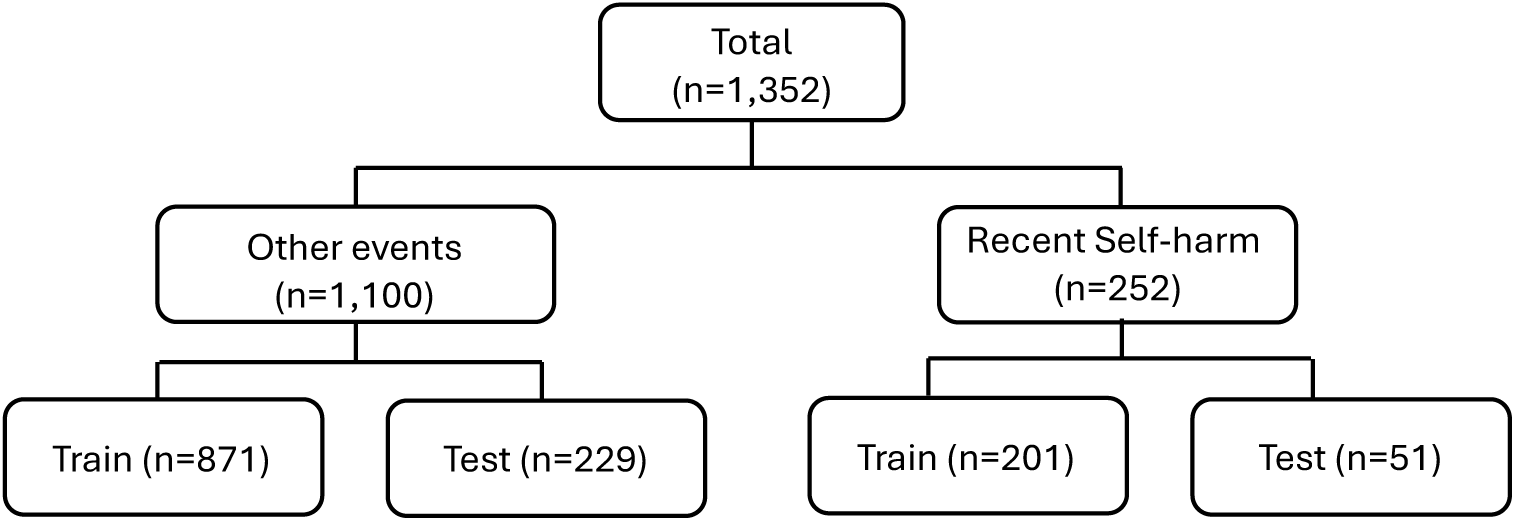
Binary relabelling of multi-label annotated examples.

We maintained identical train-test splits to ensure fair comparison between a baseline RoBERTa and Gemma3-27b models. The baseline RoBERTa model was retrained from scratch using binary cross-entropy loss optimised for the new labels. For Gemma3-27b, we retained the original multi-label prompt, then programmatically converted its structured output: cases labelled as both “self-harm present” AND “recent” were classified as “Recent self-harm”; all other label combinations mapped to “Other events.” The results are presented in Table 4.

**Table 4:**
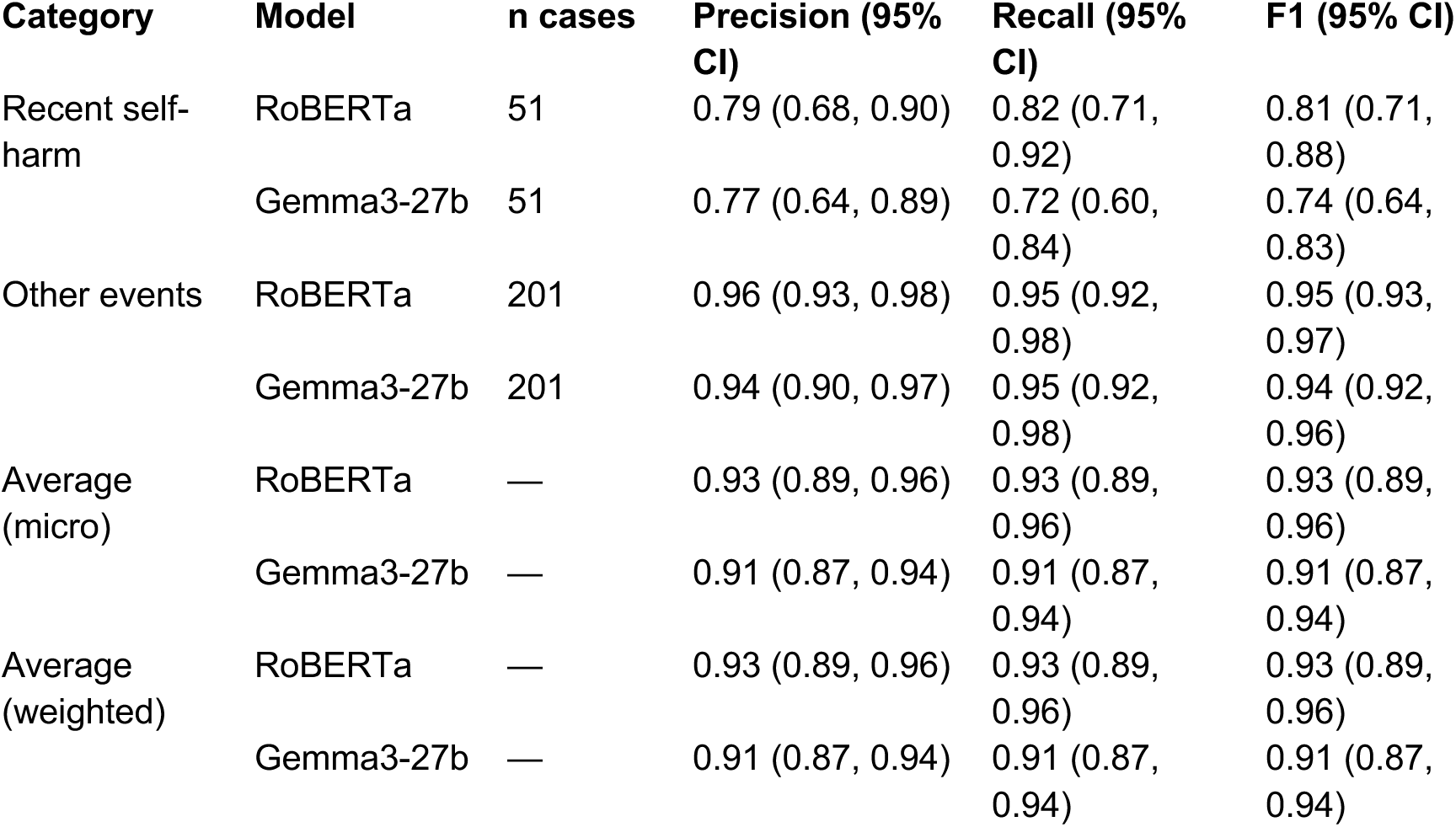
Performance of two models identifying recent self-harm. The alternative category ‘Other events’ includes any combinations, such as unconfirmed self-harm, historical events of confirmed self-harm or self-harm where timing is unknown.

The results confirmed that privacy-preserving language models can effectively identify time-sensitive clinical events. While both models demonstrated strong performance for identifying recent self-harm, the re-trained RoBERTa model achieved marginally better performance with an average weighted F1-score of 0.93 (95% CI 0.89, 0.96) compared to the F1-score for Gemma3-27b of 0.91 (95% CI 0.87, 0.94). For the dominant “Other events” category, both models performed well (F1 > 0.94) with almost identical performance.

## Discussion

### Principal findings

In this study, we aimed to evaluate whether privacy-preserving local language models could identify self-harm and its timing in secondary mental health records. The Gemma3-27b model, containing 27 billion parameters with a 128K context window, was quantised to 4-bit precision and deployed locally via Ollama, ensuring complete data privacy within the host healthcare provider’s secure data infrastructure (in our case, the National Health Service). In a corpus of 1,352 mental-health clinical notes, Gemma3-27b outperformed a fine-tuned RoBERTa classifier on both detection of self-harm events and assignment of self-harm timing labels. The absolute F1 gain was modest for event detection (≈ 4 - 6%) but substantial for challenging “historical” and “unknown” timing categories (gains of 33% and ≥ 39%, respectively). Performance on the “recent” category reached an F1 of 0.79 without gradient-based fine-tuning, although this result depended on systematic prompt development using a labelled development set. The largest relative improvements over RoBERTa were observed in the rarer timing categories, while aggregate gains were modest.

On the binary task of identifying recent self-harm, RoBERTa achieved a marginally higher weighted F1-score (0.93) than Gemma3-27b (0.91), although both models performed strongly and were comparable on the dominant ’Other events’ class. This indicates that supervised classifiers can be highly effective when the classification task is simplified and sufficient labelled data exist. The relative advantage of the prompt-based approach is most evident in the multi-label temporal setting, particularly for rarer timing categories where supervised models struggle without large per-class annotation volumes.

### Comparison with previous work

Ayre and colleagues [14] developed a hybrid rule-based NLP tool using spaCy to identify perinatal self-harm in EHRs from the South London and Maudsley NHS Foundation Trust, achieving micro-averaged F1-scores greater than 0.8 for span, polarity, and temporality detection. However, their approach required extensive manual feature engineering, custom tokenisation rules, and lexicon development. Similar to our findings, they reported temporality as the most challenging attribute (*IZ* = 0.62) and successfully employed a heuristic requiring two or more mentions for patient-level classification. While their rule-based system performed well, it required 13 manually curated lexicons and complex grammatical rules, highlighting the engineering burden of traditional NLP approaches. In contrast, our prompt-based Gemma3-27b achieved comparable or superior performance without task-specific feature engineering, demonstrating the efficiency gains of using modern LLMs. It is worth noting that both studies identified the same clinical challenge: the ambiguity in temporal expressions within clinical documentation, suggesting that this represents a fundamental limitation in how clinicians record self-harm events, rather than a purely technical challenge.

### Clinical and public health implications

Accurate ascertainment of self-harm is crucial for improving self-harm surveillance, evaluating services, and testing new interventions designed to support people who self-harm. It is also vital for identifying individuals in need of support. As the Gemma3-27b model requires relatively low computing resource, it can be deployed on in-house GPUs within a healthcare provider’s secure data infrastructure. This mitigates concerns about the use of ‘as a service’ proprietary language models hosted outside the provider’s own infrastructure where inference using prompting with patient-level data cannot be guaranteed to be consistent with relevant and territory-specific statutory regulation. The approach demonstrated here, namely locally developed and quantised language models deployed within a secure data environment, establishes the technical feasibility of privacy-preserving self-harm detection. Potential future applications include batch or near-real-time processing pipelines, clinical dashboards, and pseudonymised analytics. However, evaluation of operational feasibility, governance workflows, clinician-review safeguards, and scalability is beyond the scope of the present study and would require dedicated implementation and prospective validation studies. While prompt-based approaches may reduce, though not eliminate, the need for large volumes of annotated training data and may facilitate adaptation to related clinical tasks such as method-specific self-harm detection, suicidal ideation, or protective factors, these extensions remain speculative and require empirical validation.

### Utility and potential applications of this tool and its future iterations

Self-harm is often imprecisely captured across settings, including in the UK. This tool can support efforts to improve the monitoring of self-harm within clinical populations where such information is recorded narratively. Reliable identification and tracking of self-harm over time can provide valuable insights into temporal trends of self-harm and help assess the impact of public health policies or broader societal events [36].

Such a tool could further facilitate the identification of individuals who have recently self-harmed and may be candidates for pharmacological or psychological interventions, enabling the recruitment of representative and diverse patient samples. Moreover, given that self-harm is a key outcome in mental health care, the tool can facilitate the extraction of such information to assess the impact of targeted interventions.

Systematic and reliable identification of self-harm in EHRs is also important for estimating the burden of self-harm within clinical settings, which is essential for service planning and the allocation of resources. Similarly, it can contribute to better quality self-harm research, especially where research questions require establishing the timing of self-harm to conduct longitudinal analyses.

### Qualitative error analysis

To improve transparency about model limitations, we conducted a qualitative examination of Gemma3-27b misclassifications on the held-out test set, identifying five recurring failure modes (detailed in Appendix G with synthetic clinical-note examples constructed by the clinical team for governance compliance). These included false-positive self-harm detection, where templated risk-assessment language was mistaken for a confirmed act; false-negative detection, where self-harm described briefly within longer psychosocial narratives was overlooked; false-negative recency, where vague temporal expressions (e.g., “a few weeks ago”) were defaulted to non-recent despite falling within the 90-day window; false-positive recency, where present-tense clinical concern led the model to override explicit historical date markers; and false-positive unknown timing, where indirect but sufficient temporal cues (e.g., age-based reasoning) were not integrated. A substantial proportion of these errors arose from genuine ambiguity in clinical documentation, contexts where even expert annotators required deliberation, rather than purely technical shortcomings. These findings highlight the importance of expert review of all model outputs prior to any operational deployment, and of continuous monitoring for potential data and model drift as documentation practices, clinical populations, or language model versions evolve.

### Strengths and limitations

All computation occurred behind the Oxford Health NHS Foundation Trust firewall with no data egress, adhering to the relevant GDPR regulations and UK Data Security & Protection Toolkit standards. Two domain experts produced a high-quality gold standard with very good κ = 0.86 for event detection. The study used identical splits and evaluation metrics for both the Gemma3-27b language model and the RoBERTa model, isolating the effect of model architecture. A single, relatively low-cost NC-T4 node (16 GB GPU) demonstrates broad feasibility across publicly funded healthcare settings, such as the NHS in the UK, without requiring large-scale high performance computing infrastructure and a low carbon footprint.

This study has six main limitations. First, while demonstrating feasibility of privacy-preserving language models for self-harm detection, we acknowledge limited model selection. We evaluated locally deployable models via Ollama (Llama 3.2, Mistral, Phi-4, Qwen 2, various Gemma3 variants), finding performance correlated with parameter count, consistent with established scaling laws [37, 38]. Optimal prompts varied substantially across models, reflecting differences in pre-training, corpora and architecture [39]. This model-specific sensitivity suggests Gemma3-27b may not represent optimal performance. We selected Gemma3-27b pragmatically as a representative high-performing model, balancing computational resources with demonstrating feasibility rather than identifying the optimal clinical deployment model. When using privacy-preserving, local quantised LLMs in applications similar to those in our study, it will be important to systematically evaluate different model architectures and prompting strategies.

Second, data for training and testing the models were sourced from a single region and secondary mental healthcare setting in England; therefore, external validity to other regions with different populations, or to primary and acute care settings, was not assessed. Of note, the nature of underlying data differs substantially between secondary mental health care (the data used in our study) and primary or acute care settings due to variations in clinical practice. For example, acute hospitals rely more heavily on structured clinical coding to record patient presentations and encounters involving self-harm, whereas mental health care data place greater emphasis on narrative psychosocial formulations of historical and current self-harm and its management. For these reasons, we would expect that, across the UK’s secondary mental health care system, the presented model would show limited variation in performance for the self-harm task described here, as services share a similar culture of practice and use EHRs with comparable functionality. However, a substantially different model would likely be required to address the same task in acute or primary care EHRs.

Third, a potential selection effect arises from using a Gemma3-4b model for candidate-note identification and a Gemma3-27b model as the primary evaluation model. Although these models differ substantially in parameter count (4 billion vs. 27 billion) and the screening step was used solely to make annotation feasible, it did not generate gold-standard labels, and no model parameters were updated based on screening outputs, both models belong to the same architectural family. If Gemma-family models share systematic biases in what they flag as self-harm-related, the evaluation corpus could, in principle, contain a distributional signature that favours Gemma3-27b over architecturally different models such as RoBERTa. Two observations mitigate this concern: (a) the screening step removed many “easy negatives” (e.g., administrative notes), yielding a harder evaluation set containing more ambiguous cases, which are consistent with the study’s clinical aims; and (b) RoBERTa achieved strong overall performance and marginally outperformed Gemma3-27b on the simplified binary task, which would not be expected under strong architectural bias. Nevertheless, a residual distributional effect cannot be excluded. Future validation should include sensitivity analyses, such as alternative screening strategies (e.g., keyword-based or clinician-led) or a supplementary truly random annotated sample, to quantify the magnitude of any selection effect.

Fourth, because the annotated corpus is enriched for potential self-harm content, the class distribution does not reflect the prevalence that would be encountered when the model is deployed across all clinical notes in routine care. In a low-prevalence setting, even a model with high specificity can generate a non-trivial number of false positives at scale, increasing clinician review burden and potentially undermining trust. Prospective evaluation under true-prevalence conditions, prevalence-aware calibration or thresholding strategies, and clinician-in-the-loop workflows in which every model output is reviewed before any clinical action, are essential prerequisites for operational deployment.

Fifth, this study did not evaluate operational deployment considerations. Questions relating to batch versus near-real-time processing architectures, governance frameworks for automated flagging, human-in-the-loop safeguards, and real-world performance under routine clinical conditions were beyond the scope of the present work and constitute essential future research.

Sixth, many patients had a long-documented history of contact with secondary mental health services. Although annotators were instructed to treat each clinical note as a standalone document, independent of any previous decisions made about previous extracts from the same patient, this may not have been fully achievable in practice. As a result, some annotations may have been influenced by broader impressions of the patient rather than by information explicitly present in the text being analysed. Historical (90 of 1,352, 6.7%) and unknown (77 of 1,352, 5.7%) timing cases were under-represented, inflating confidence intervals despite bootstrap resampling. Future work should use active-learning strategies to enrich rare labels. The 90 days threshold, while pragmatic, may not entirely align with all clinical use-cases; finer-grained temporal consensus on recency remains challenging. However, the recency threshold could be readily changed by amending the prompt.

## Conclusion

This work demonstrates the technical feasibility of using a privacy-preserving, locally deployable language model within secure NHS data infrastructure to identify self-harm and its timing. Without gradient-based fine-tuning, but with systematic prompt development on a labelled development set, Gemma3-27b matched or exceeded a fine-tuned RoBERTa classifier, with the largest gains in challenging, lower-frequency timing categories and modest aggregate improvements. On a simplified binary task, RoBERTa performed marginally better, highlighting that the choice of approach should be guided by the specific clinical task and available annotation resources. These findings establish a proof of concept; clinical deployment would require multi-centre validation across geographically diverse areas and populations, prospective evaluation of operational workflows (including clinician-review safeguards, governance frameworks, and false-positive management), implementation research with stakeholders, and rigorous monitoring for model drift and unintended bias. Such studies are the critical next steps to translate privacy-preserving language models into improved self-harm surveillance and patient confidentiality.

## Data Availability

The data used in this work are owned by Oxford Health NHS Foundation Trust and accessed throught CRIS Powered by Akrivia Health, using anonymised patient records. The data cannot be made publicly available but can be accessed with permissions from Oxford Health NHS Foundation Trust for UK NHS staff and UK academics within a secure firewall, in the same manner as the authors.

## Data availability

The data used in this work are owned by Oxford Health NHS Foundation Trust and accessed through CRIS Powered by Akrivia Health, using anonymised patient records. The data cannot be made publicly available but can be accessed with permissions from Oxford Health NHS Foundation Trust for UK NHS staff and UK academics within a secure firewall, in the same manner as the authors.

## Acknowledgements

This research is funded by NIHR Oxford Health Biomedical Research Centre (NIHR203316) and supported by Oxford Health NHS Foundation Trust Research Informatics Team. The study was carried out using OHFT electronic patient records within the Akrivia CRIS research platform environment, owned by Akrivia Health (akriviahealth.com).

GG and RB were supported by funding from Australia’s National Health and Medical Research Council (NHMRC) awarded to RB (#GNT2008073). GG was supported in part by the Department of Health and Social Care through a grant for the Multicentre Study of Self-harm in England.

AK and DWJ were supported in part by the NIHR AI Award for Health and Social Care (AI_AWARD02183), AK by a research grant from GlaxoSmithKline. This study was supported by CRIS Powered by Akrivia Health, using data, systems and support from the NIHR Oxford Health Biomedical Research Centre (BRC-1215-20005) Research Informatics Team.

DWJ was in-part supported by the Office for Life Sciences and the National Institute for Health and Care Research (NIHR) Mental Health Translational Research Collaboration Mission, hosted by the NIHR Oxford Health Biomedical Research Centre.

NK is supported by the National Institute for Health and Care Research Greater Manchester Patient Safety Research Collaboration (NIHR204295), the University of Manchester and Mersey Care NHS Foundation Trust.

We would like to thank the collaborators on the NIHR-ARC application, Professor Keith Hawton, Professor Andrea Cipriani, and Professor Seena Fazel, as well as Liz Bale for her contribution to annotating the data. We would also like to acknowledge the work and support of the Oxford Research Informatics Team for their support throughout this project: Adam Pill, Acting Joint Head of Research Informatics, Suzanne Fisher, Research Informatics Systems Analyst, Lulu Kane, Research Informatics Administrator and Tanya Smith.

The views expressed are those of the authors and not necessarily those of the UK National Health Service, the NIHR or the UK Department of Health and Social Care.

## Authors’ Contributions

Conceptualisation: G.G., A.K., A.T.; Methodology: G.G., A.K.; Data acquisition: G.G., A.K., A.T.; Formal analysis: G.G., A.K.; Draft manuscript: G.G., A.K.; Review and editing: G.G., A.K., A.T., R.B., D.J., N.K. All authors have read and agreed to the published version of the manuscript.

## Appendix

### A. Cohort statistics

**Table A.1.** The annotated cohort description used to develop and validate models.

| Characteristic | n (%) or median [IQR] |
| --- | --- |
| **Demographics** |  |
| Total clinical notes | 1,352 |
| Total unique patients | 628 |
| Age<sup>b</sup>, years | 37 [25.8, 53.3] |
| 18–24 | 151 (24) |
| 25–44 | 233 (37.1) |
| 45–64 | 191 (30.4) |
| ≥65 | 53 (8.4) |
| **Sex** |  |
| Female | 355 (56.5) |
| Male | 273 (43.4) |
| **Ethnicity** |  |
| White | 463 (84.2) |
| Asian | 23 (4.2) |
| Mixed | 20 (3.7) |
| Black | 11 (2.0) |
| Other | 5 (1.7) |
| Not stated/Unknown | 106 (16.9) |
| **Top 10 Primary Mental Health Diagnoses (ICD-10)<sup>a</sup>** |  |
| **Psychotic disorders (F20–F29)** |  |
| F20 – Schizophrenia | 45 (9.9) |
| F25 – Schizoaffective disorders | 19 (3.8) |
| **Mood disorders (F30–F39)** |  |
| F31 – Bipolar affective disorder | 47 (9.5) |
| F32 – Depressive episode | 89 (17.9) |
| F33 – Recurrent depressive disorder | 34 (6.9) |
| **Neurotic/stress-related (F40–F48)** |  |
| F41 – Other anxiety disorders | 31 (6.3) |
| F43 – Reaction to severe stress | 22 (4.4) |
| **Behavioural syndromes (F50–F59)** |  |
| F50 – Eating disorders | 16 (3.2) |
| **Personality disorders (F60–F69)** |  |
| F60 – Specific personality disorders | 36 (7.3) |
| **Self-harm History (from notes analysed)** |  |
| Any self-harm mentioned | 419 (30.9) |
| Recent (≤90 days) | 252 (18.6) |
| Historical (>90 days) | 90 (6.6) |
| Timing unclear | 77 (6) |
| **Note Characteristics** |  |
| Notes per patient, median [IQR] | 2 [1, 3] |
| Note length (words), median [IQR] | 248 [146, 315] |

IQR = interquartile range.
Patients may have multiple diagnoses.
a Percentages based on unique patients (n=628).
b Age at first admission to secondary mental health care services.

### B. Selection of clinical notes that may contain mentions of self-harm

We used a light-weight version of ‘Gemma3’ model with 4 billion parameters to pre-select clinical notes that may contain mentions of self-harm without any further differentiation. The model was served via Ollama (0.9.6) framework using Python (3.12.8). The internal parameters of ‘Gemma3-4b’ were: temperature = 0, top_p = 0.95, top_k=64. Average end-to-end latency was 1.5 s per 500-token note on a single Microsoft Azure ‘Standard_NC8as_T4_v3’ instances (NVIDIA T4 16GB).

The following is the verbatim prompt used to pre-select clinical notes in the study; it is included for reproducibility and does not contain patient data.

###################### BEGINNING OF THE PROMPT ###################### ### SYSTEM

You are an expert clinical-text classifier.

Your only job is to decide whether the note you receive contains **any explicit or implicit mention of self-harm** (suicidal ideation, self-injury, suicide attempts, self-poisoning, thoughts of wanting to die, etc.).

If the note contains even one such reference, answer **“YES”**.

If it does **not** (e.g., routine referrals, appointment letters, discharge summaries without self-harm), answer **“NO”**.

\### OUTPUT RULES

1. Output **only** a single JSON object that exactly matches this schema:

{

“self_harm”: “YES” | “NO”, “reasoning”: “string”

}

\### DEFINITIONS

– self_harm must be either “YES” or “NO” (uppercase).

– reasoning is a concise phrase up to 100 words justifying the label; do not expose chain-of-thought.

2. No additional keys, comments, markdown, or extra text, just the JSON.

\### DECISION GUIDELINES

Label YES if the note includes any of the following (even once):

- Suicidal thoughts, plans or attempts

- Non-suicidal self-injury (cutting, burning, overdosing)

- Historical self-harm that is clinically relevant to the note

- Risk assessments that state self-harm risk/ideation is present

Label NO when:

- The note is administrative (appointment, referral, billing)

- It is a clinical summary with no self-harm content

- Self-harm is explicitly denied and no other self-harm content appears

- Edge cases then err on the side of YES.

\### EXAMPLES INPUT NOTE A

“Patient reports cutting her forearm three days ago after argument with partner.” OUTPUT

{“self_harm”:“YES”, “reasoning”:“Recent self-injury (cutting) described.”}

INPUT NOTE B

“Follow-up letter confirming physiotherapy appointment for lower-back pain.” OUTPUT

{“self_harm”:“NO”, “reasoning”:“Administrative appointment; no self-harm content.”}

\### TASK

Classify the following clinical note:

{note}

######################### END OF THE PROMPT #########################

### C. Development of the self-harm detection prompt

We experimented with several approaches to capture nuanced information about self-harm episodes and their temporal characteristics. The development followed systematic design patterns recommended for distilled language models like Gemma3-27b. We developed comprehensive exclusion criteria through negative examples taken from clinical notes, including thoughts, plans, threats, and acts by others. Additionally, we included specific guidance for handling ambiguous scenarios and edge cases.

The initial prompt contained extensive instructions with multiple restatements of key concepts. While thorough, this approach suffered from several limitations: (i) lengthy (∼1,200 words) with repeated instructions; (ii) scattered organisation lacking logical flow; (iii) inconsistent emphasis patterns, and (iv) absence of concrete examples.

We, therefore revised our approach and applied structured prompting principles to improve the prompt by introducing clear section headers for better navigation, grouped related instructions for coherence, paraphrasing repeated structures, added examples and precedence rules for disambiguation of multiple incidents (e.g., recent versus historical). In the final version, we expressed the temporal boundary as a precise 90-day threshold using mathematical notation. We introduced formal notation such as “RD” for record date to enhance clarity. Further, we adopted the following naming convention (e.g., “NOSH” stand for “<u>NO S</u>elf-<u>H</u>arm”), following Table C.1:

**Table C.1:**
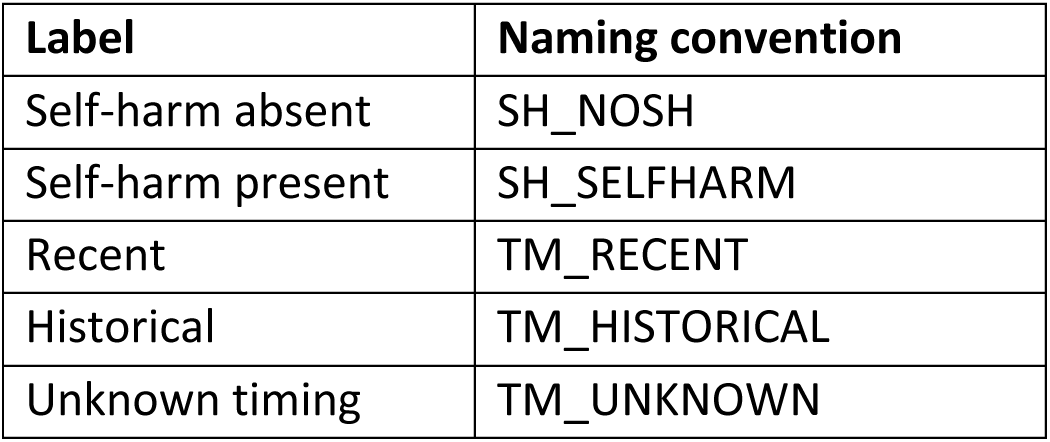
Labelling convention used in this study.

| Label | Naming convention |
| --- | --- |
| Self-harm absent | SH_NOSH |
| Self-harm present | SH_SELFHARM |
| Recent | TM_RECENT |
| Historical | TM_HISTORICAL |
| Unknown timing | TM_UNKNOWN |

We strictly instructed the model to generate output in JSON format to ensure consistency, enhanced by using the Python’s ‘pydantic’ class. The final prompt was reviewed by two experts in self-harm for its clinical correctness and consistency. To ensure reproducibility, the temperature parameter of the Gemma3-27b model was set to zero. All examples shown in the prompt below are made up by the clinical team according to data governance. Below is the final prompt used in this work.

The following is the verbatim prompt used in the study; it is included for reproducibility and does not contain patient data.

###################### BEGINNING OF THE PROMPT ######################

\### SYSTEM

You are a clinical-NLP assistant analysing patient records for self-harm.

\### TASK

For the clinical note below (starts with [YYYY-MM-DD]), determine:

1. Did the patient perform an intentional act of self-injury or self-poisoning?
2. If yes, when did it occur?

\### DEFINITIONS

Self-harm = Any COMPLETED intentional act of self-injury or self-poisoning by the patient, regardless of method or motive.

\### CLASSIFICATION RULES

Label as SH_SELFHARM only if:

- Clear evidence of a completed act (e.g., “cut wrist”, “took 20 tablets”, “burned with cigarette”)

- Conflicting information: If act clearly occurred, classify as SH_SELFHARM even if later denied

Label as SH_NOSH if:

- Thoughts, ideation, plans, or threats without action

- Risk assessments or statements about potential

- Acts by others (family, friends)

- Preparations without execution (e.g., “held pills but didn’t take”)

- No clear evidence of completed act

RECENCY (only if SH_SELFHARM)

Let RD = record date at start of note.

- TM_RECENT: Act occurred ≤90 days before RD OR uses phrases like “yesterday/last week/past month” OR describes ongoing behaviour

- TM_HISTORICAL: Act occurred >90 days before RD OR uses phrases like “last year/as a teenager”

- TM_UNKNOWN: Timing cannot be determined

- Priority: If multiple acts with different timing, return TM_RECENT

\### OUTPUT

Return ONLY this JSON (no markdown, no explanations):

{

“self_harm”: “SH_SELFHARM” or “SH_NOSH”,

“recency”: “TM_RECENT” or “TM_HISTORICAL” or “TM_UNKNOWN”,

“method”: “” or “Unknown”, “evidence”: “” or “Unknown”,

“self_harm_date”: “” or “Unknown”, “record_date”: “”

}

\### EXAMPLE

Input: [2024-03-15] “Patient reports she cut her arms with a razor blade last Tuesday…” Output:

{

“self_harm”: “SH_SELFHARM”, “recency”: “TM_RECENT”,

“method”: “cutting with razor blade”,

“evidence”: “cut her arms with a razor blade last Tuesday”, “self_harm_date”: “last Tuesday”,

“record_date”: “2024-03-15”

}

Analyse this note:

{text}

######################### END OF THE PROMPT #########################

### D. Model performance

Below are the multi-label confusion matrices of RoBERTa (n=1,084) (left column) and Gemma3-27b (right column) for each of the categories. Since presence and absence of self-harm are exclusive categories, both labels can be presented by a single matrix.

**Figure D.1.**
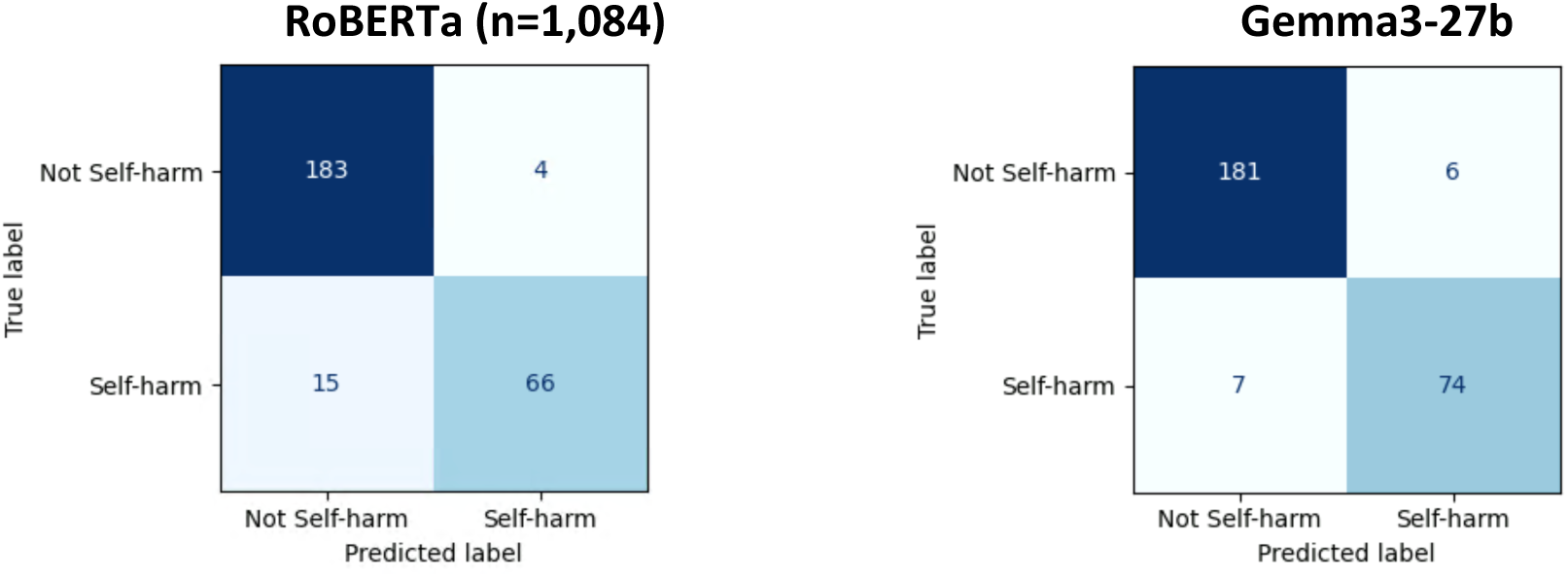
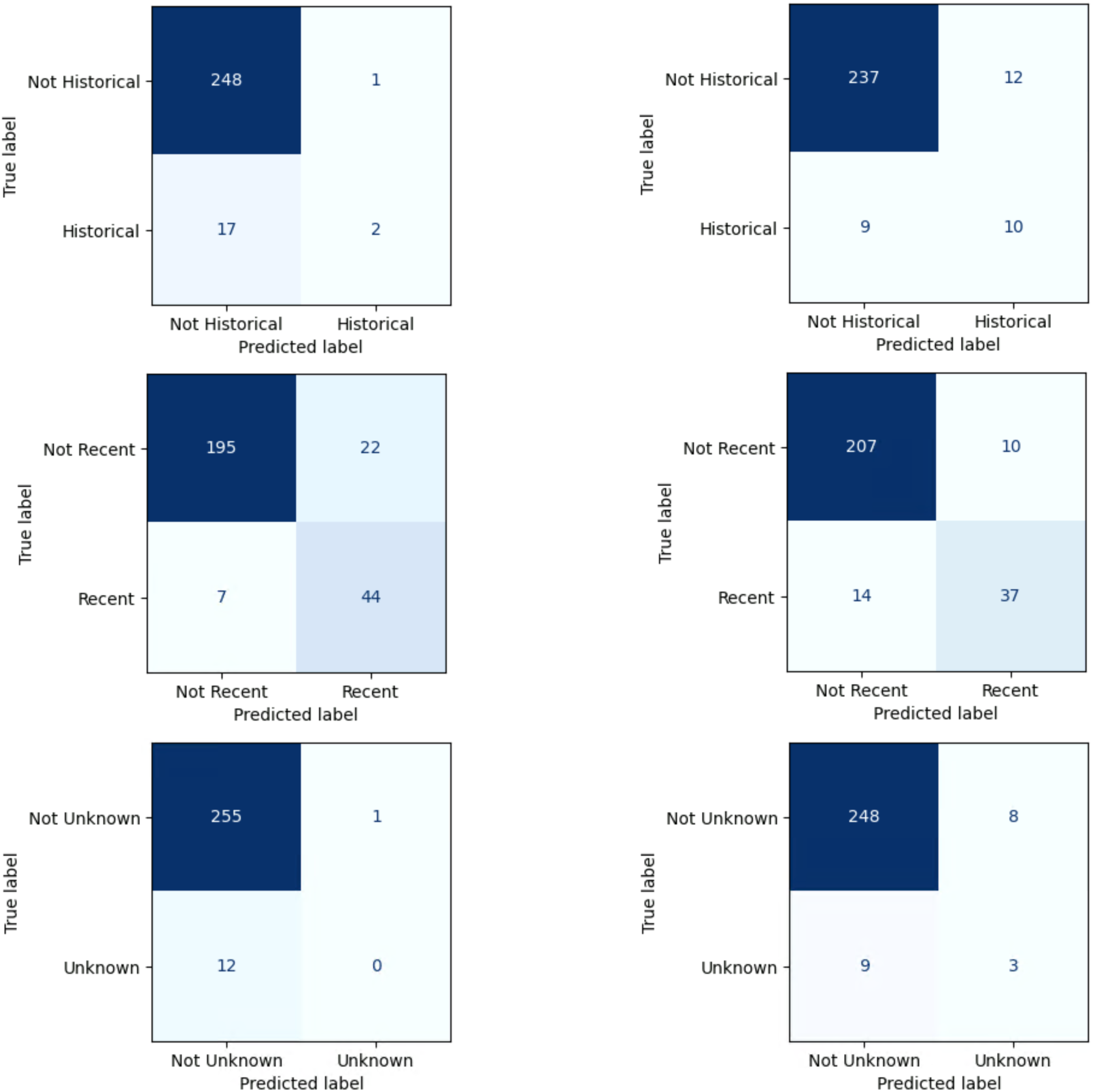
Comparison of multi-label confusion matrices for each of the categories.

Each matrix shows the performance of one specific label independently, showing how well the model predicts that particular (i.e., positive class) label. In these matrices, each label is treated as a binary classification problem, therefore, the “label” represents instances where the positive (predicted) class is present, while “not label” represents instances where that specific label is absent. Across categories, Gemma3-27b appears to be more balanced between false positive and false negative predictions, while RoBERTa captures more true positives as well as more false positives.

### E. Statistical significance testing of model performance

Here we present detailed statistical comparisons between RoBERTa(n=1,084) and Gemma3-27b models using two-sided McNemar’s test for multi-label classifications and bootstrap analysis of F1 scores. All tests were performed on the held-out test set n=268 with 10,000 bootstrap iterations with bias-corrected percentile intervals. Multiple comparison correction was applied using the Benjamini-Hochberg false discovery rate (FDR) method with α=0.05.

Binarisation of Multi-label Predictions

Since McNemar’s test requires binary paired outcomes, we transformed our multi-label predictions using a one-vs-rest approach. For each of the five labels, we created binary predictions using the following algorithm (Table E.1):

**Table E.1:** Binarisation of multi-label schema for application of the McNemar’s test.

|  |  |
| --- | --- |
| Self-harm absent | 1 if predicted as absent, 0 otherwise |
| Self-harm present | 1 if predicted as present (regardless of timing), 0 otherwise |
| Recent | 1 if predicted as self-harm present AND recent, 0 otherwise |
| Historical | 1 if predicted as self-harm present AND historical, 0 otherwise |
| Unknown timing | 1 if predicted as self-harm present AND unknown timing, 0 otherwise |

Each McNemar’s test compared these binarised predictions between models for a single label. The p-values in Table E.2 represent five independent tests, which is why we applied Benjamini-Hochberg correction for five comparisons (not multiple tests per label).

**Table E.2:**
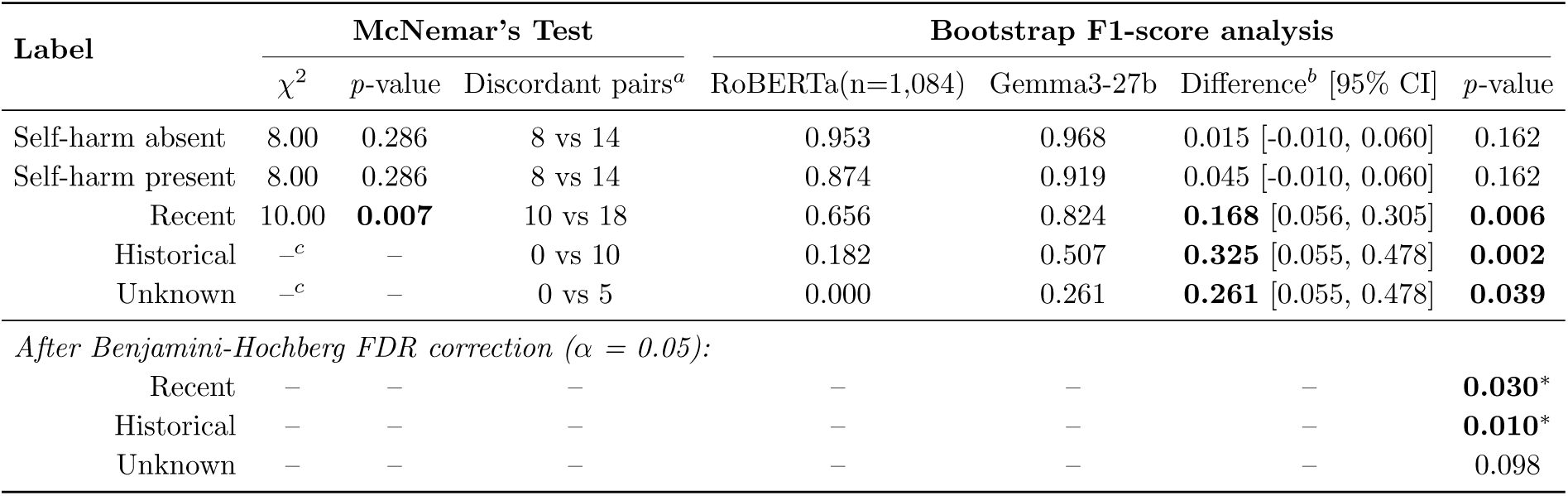
Statistical comparison of RoBERTa(n=1,084) vs Gemma3-27b performance using McNemar’s test and bootstrap analysis. ^a^ Number of cases where only RoBERTa vs only Gemma3-27b correctly predicted this label (binarised). ^b^ Difference = Gemma3-27b F1 - RoBERTa F1, calculated on binarised predictions for each label. ^c^ McNemar’s test undefined when one model has zero unique correct predictions for this label. ^d^ Each row represents an independent McNemar’s test on binarised predictions for that specific label. * Remains significant after FDR correction for 5 independent comparisons (one per label), bold indicates statistical significance at α=0.05.

The statistical analysis showed that Gemma3-27b significantly outperforms RoBERTa(n=1,084) on the most clinically challenging temporal classification tasks. McNemar’s test showed significant differences for recent self-harm detection (χ²=10.00, p=0.007), with Gemma3-27b correctly identifying 18 cases missed by RoBERTa(n=1,084), while RoBERTa(n=1,084) uniquely identified only 10 cases. Bootstrap analysis with 10,000 iterations confirmed these findings and demonstrated the performance gains: (i) recent self-harm:

Gemma3-27b achieved 16.8% higher F1 score (95% CI: 5.6-30.5%, p=0.006), (ii) historical self-harm: 32.5% improvement (95% CI: 5.5-47.8%, p=0.002) and (iii) unknown timing: 26.1% improvement (95% CI: 5.5-47.8%, p=0.039).

After Benjamini-Hochberg correction for multiple comparisons, the differences for recent (adjusted p=0.03) and historical (adjusted p=0.01) classifications remained statistically significant, while unknown timing became marginally non-significant (adjusted p=0.098).

RoBERTa(n=1,084) underperformed entirely to identify any historical or unknown timing cases that Gemma3-27b missed (0 unique correct predictions), demonstrating the supervised model inability to learn from rare categories despite training on the full dataset. This demonstrates the additional value of large language models pre-trained knowledge for handling clinical edge cases where training examples are scarce.

### F. Reproducibility and details of models training

All experiments were conducted on Microsoft Azure ‘Standard_NC8as_T4_v3’ instances (NVIDIA T4 16GB). The baseline RoBERTa model, implemented in PyTorch (v2.4.1) using HuggingFace library (v4.54.0), was fine-tuned for multi-label classification using binary cross-entropy loss with logits. The base RoBERTa architecture contains 12 transformer layers, 768 hidden dimensions, and 12 attention heads (125M parameters). Training used AdamW (learning rate of 2 × 10⁻⁵, β₁ = 0.9, β₂ = 0.98), linear warm-up over 500 steps with batch size = 8 for 10 epochs. One NVIDIA T4 16GB completed 10 epochs under 20 min.

The Gemma3-27b model, a decoder-only transformer containing 27 billion parameters with a 128K token context window, was quantized to 4-bit precision (Q4_K_M format) and converted to GPT-Generated Unified Format (GGUF), resulting in a 10.6 GB model size. The model employs a novel 5:1 local-to-global attention layer architecture with 1,024-token sliding windows for memory-efficient inference. It was served locally via the Ollama framework (v0.9.6) with llama.cpp backend. No gradient-based training was performed; instead, the model relied on zero-shot prompting with the carefully engineered prompt described in Appendix C.

For our experiments, we configured the context window to 8,192 tokens to balance performance and computational resources. Deterministic decoding (temperature=0, top_p=0.95 and top_k=64) produced the required JSON output which was parsed to extract labels. Average end-to-end latency was 90 seconds per 500-token clinical note on a single NVIDIA T4 16GB GPU served via Microsoft Azure.

All codes, annotated data and trained models can be made available upon request for users that are authorised on the Akrivia Health research platform.

### G. Qualitative analysis of failure modes

To improve transparency about model limitations, we examined the major recurring patterns of misclassification (“failure modes”) produced by Gemma3-27b on the held-out test set. For each failure mode we describe the common pattern and provide a synthetic clinical note excerpt that illustrates the type of language involved. All examples below were constructed by the clinical team and do not reproduce patient text.

1. False-positive self-harm - the model predicted a self-harm event where the gold standard confirmed that no actual act took place.

Common pattern: The model interpreted risk-assessment language, documentation of self-harm ideation, or references to historical self-harm within templated safety-plan fields as evidence of a completed act. Clinical notes that discuss self-harm potential or past self-harm purely in the context of ongoing risk monitoring, without describing a new or specific act, were particularly prone to this error.

Synthetic example: *[2023-06-14] Review in community team. Risk assessment updated. History of self-harm by overdose, last known episode approximately two years ago. Current risk: moderate. Patient denies any recent self-harm or suicidal ideation. Plan: continue fortnightly contact, crisis plan reviewed*.

Here, the note documents historical self-harm within a risk assessment, but explicitly states there has been no recent act. The model incorrectly classified this as self-harm present, apparently triggered by the phrase “self-harm by overdose” despite the negating context.

2. False-negative self-harm - the model missed a confirmed self-harm event by predicting its absence.

Common pattern: The model failed to recognise self-harm when the clinical language was indirect, euphemistic, or embedded within a longer narrative about the patient’s psychosocial circumstances. Notes where the self-harm act was described briefly alongside extensive discussion of stressors, mental state, or treatment plans were particularly affected, as though the surrounding context diluted the salience of the event.

Synthetic example: *[2024-01-22] Home visit. Patient appeared low in mood, described ongoing difficulties with housing and finances. Mentioned she had taken a handful of her prescribed tablets last Thursday following an argument, felt unwell but did not seek medical attention. Discussed safety planning and coping strategies. Will refer to psychology*.

The note contains a clear self-harm act (“taken a handful of her prescribed tablets”), but the model classified it as self-harm absent, apparently because the overdose was mentioned briefly within a longer passage focused on social stressors and care planning.

3. False-negative recency - a recent self-harm event was misclassified as historical or of unknown timing.

Common pattern: When clinical text used vague or relative temporal language, such as “a few weeks ago,” “recently,” or “not long ago”, without an explicit date, the model tended to default to a non-recent classification. The absence of a precise date appeared to be treated as evidence of temporal distance rather than as insufficient information. This was most pronounced in notes where the temporal expression was syntactically separated from the description of the act.

Synthetic example: *[2024-05-10] Outpatient review. Patient disclosed that she had cut her arms a few weeks ago during a period of distress. Wounds have healed. Currently feels more stable. No active suicidal ideation*.

The self-harm occurred “a few weeks ago” relative to a note dated 10 May 2024, placing it well within the 90-day recency window. The model classified this as historical, apparently interpreting the past tense and the healed wounds as markers of a remote event.

4. False-positive recency - a historical or unknown-timing event was misclassified as recent self-harm.

Common pattern: The model incorrectly assigned recent timing when a note described current suicidal ideation, current emotional distress, or active clinical risk alongside a self-harm event that was explicitly dated beyond the 90-day window. The model appeared to apply a proximity heuristic: when any self-harm-related content co-occurred with present-tense clinical concern, it defaulted to “recent” regardless of the temporal markers attached to the act itself.

Synthetic example: *[2024-09-03] CPA review. Patient reports ongoing low mood and fleeting thoughts of self-harm. Previously took an overdose of paracetamol in January 2024, requiring A&E attendance. Currently engaging with crisis team. Risk assessment updated*.

The overdose is explicitly dated to January 2024, more than seven months before the note, and should be classified as historical. The model classified it as recent, apparently influenced by the present-tense language about ongoing ideation and crisis-team involvement.

5. False-positive unknown timing - a self-harm event with determinable timing (recent or historical) was misclassified as unknown timing.

Common pattern: The model defaulted to “unknown timing” when the temporal information, although present, was conveyed indirectly or required inference from multiple cues rather than a single explicit date. Notes containing phrases such as “last year” or “when she was a teenager”, which provide sufficient information to classify an event as historical, were sometimes labelled as unknown, suggesting the model applied an overly conservative threshold for temporal certainty rather than integrating contextual evidence.

Synthetic example: *[2024-07-18] Assessment. Patient reports that she used to cut herself regularly as a teenager; stopped around age 19. She is now 34. Scars visible on both forearms. No self-harm for many years*.

The note establishes that self-harm occurred approximately 15 years ago, clearly historical. The model classified the timing as unknown, apparently because no specific date was given, despite the age-based temporal reasoning providing sufficient information.

These failure patterns highlight that a substantial proportion of misclassifications arise from genuine ambiguity or indirectness in clinical documentation, contexts where even expert annotators required deliberation. The findings highlight the importance of expert review of all model outputs prior to any operational deployment, and of continuous monitoring for potential data and model drift as documentation practices, clinical populations, or language model versions evolve.

